# Emergence of SARS-CoV-2 variant B.1.575.2 containing the E484K mutation in the spike protein in Pamplona (Spain) May-June 2021

**DOI:** 10.1101/2021.08.11.21261950

**Authors:** Camino Trobajo-Sanmartín, Ana Miqueleiz, María Eugenia Portillo, Miguel Fernández-Huerta, Ana Navascués, Pilar Sola Sara, Paula López Moreno, Gonzalo R Ordoñez, Jesús Castilla, Carmen Ezpeleta

**Author notes:** Corresponding author: Camino Trobajo-Sanmartín.

## Abstract

With the emergence of new SARS-CoV-2 variants and the acquisition of novel mutations in exiting lineages, the need to implement methods capable of monitoring viral dynamics arises. We report the emergence and spread of a new SARS-CoV-2 variant within B.1.575 lineage containing the E484K mutation in the spike protein (named B.1.575.2) in a region of Northern Spain between May and June 2021. SARS-CoV-2 positive samples with cycle threshold value less than or equal to 30 were selected to screen of presumptive variants using the TaqPath™ COVID-19 RT-PCR kit and TaqMan™ SARS-CoV-2 Mutation Panel. Confirmation of variants was performed by whole genome sequencing. Of the 200 samples belonging to the B.1.575 lineage, 194 (97%) corresponded to the B.1.575.2 sub-lineage, which was related to the presence of the E484K mutation. Of 197 cases registered in GISAID EpiCoV database as lineage B.1.575.2 194 (99.5%) were identified in Pamplona (Spain).

This report emphasizes the importance of complementing surveillance of SARS-CoV-2 with sequencing for the rapid control of emerging viral variants.

## Introduction

During the severe acute respiratory syndrome coronavirus 2 (SARS-CoV-2) pandemic, several variants were catalogued as variants of concern (VOC) and variants of interest (VOI) by the European Centre for Disease Prevention and Control has emerged in different countries. As of June 23, 2021, the four important lineages with evidenced impact on transmissibility, severity and immunity are linage B.1.1.7 (Alpha), B.1.351 (Beta), B.1.617.2 (Delta), and P.1 (Gamma) (1-4). Lineages B.1.351 and P.1 were of specific concern because they present the spike mutation E484K, which has been associated with the reduced neutralizing activity of antibodies and may be associated with the reduced efficacy of the vaccine (2,3,5,6). Initially, the B.1.1.7 lineage had mutations N501Y and D614G and the characteristic ΔH69/ΔV70 deletion in the spike protein; however, in early 2021, Public Health England reported the first B.1.1.7 SARS-CoV-2 cases that had acquired the E484K mutation (3,7).

In this regard, concerns about the emergence of new SARS-CoV-2 variants and the acquisition of new mutations in existing lineages, such as the accumulation of mutations in the spike gene in B.1.1.7, have been developing since the onset of the pandemic. This study identified the emergence and spread of the E484K spike mutation within the SARS-CoV-2 B.1.575 lineage that has increased in the circulating virus population in Pamplona (Spain) between May and June 2021. Additionally, we share our experience with the prospective surveillance of novel SARS-CoV-2 variants by implementing a two-step laboratory strategy: reverse transcription quantitative real-time polymerase chain reaction PCR (RT-qPCR) screening and whole genome sequencing.

## Materials and methods

The Microbiology Department of the Complejo Hospitalario de Navarra, located in Pamplona, the capital city of Navarra (Spain), is the reference laboratory of the public health system for SARS-CoV-2 (approximately 650,000 inhabitants). Upper respiratory specimens for SARS-CoV-2 detection are routinely collected at the hospital and primary care centers, and processed by commercial RT-qPCR methods. Since the end of 2020, when variant B.1.1.7 became predominant in the United Kingdom, prospective sample-based surveillance has been conducted in our community to identify novel emerging SARS-CoV-2 variants. A two-step laboratory procedure included all positive SARS-CoV-2 samples from hospital patients and community settings with a cycle threshold (Ct) less or equal to 30. Occasionally, targeted samples are also included according to epidemiological criteria.

Screening of presumptive SARS-CoV-2 variants carrying ΔH69-ΔV70 deletion was performed using the TaqPath™ COVID-19 RT-PCR kit (Thermo Fisher Scientific, USA) following the manufacturer’s instructions. Then, all those samples non-B.1.1.7 variants were subjected to a second RT-qPCR, TaqMan™ SARS-CoV-2 Mutation Panel (Thermo Fisher Scientific, USA). At that moment, we customed TaqMan assay for detecting SARS-CoV-2 spike protein with the N501Y, E484K, K417N, K417T mutations. All samples were sequenced.

Whole-genome sequencing was performed using Illumina COVIDSeq Test (Illumina Inc, USA) on the Illumina NovaSeq 6000 located in the public company NASERTIC, following the manufacturer’s instructions. The viral lineages classifications were performed by the Global Initiative on Sharing Avian Influenza Data (GISAID) (https://www.gisaid.org/) [GISAID EpiCoV] database, Nextstrain (https://nextstrain.org/) [Nextstrain] and Phylogenetic Assignment of Named Global Outbreak (PANGO) Lineages (https://cov-lineages.org/) [Pango] (8-10).

## Results

As of August 1, 2021, a total of 4,728 SARS-COV-2 genomes have been sequenced in Navarra. Our sequencing analysis of the SARS-CoV-2 identified 200 (4.2%) samples related to the B.1.575 lineage: four (2%) B.1.575, two (1%) B.1.575.1 and 194 (97%) B.1.575.2. Among the common substitutions present in these lineages, four occurred in the spike protein (S494P, D614G, P681H, T716I). (11). All samples showed a gene S positive (not carrying ΔH69-ΔV70 deletion) in TaqPath. In TaqMan, all samples identified by sequencing as B.1.575.2 showed the E484K mutation.

The first case with B.1.575 lineage to be identified in Pamplona dates back to January 20, 2021; since that date, no other case was identified until March 15, 2021, where three isolates showing mutations common to the B.1.575 lineage were recorded.

Between weeks 20 to 26 2021, we identified 194 cases with lineage B.1.575, which had acquired another S mutation, E484K, classified in the GISAID EpiCoV and Pangolin databases as the sub-lineage B.1.575.2. The first case with B.1.575.2 lineage was identified in a sample isolated on May 19 (week 20, 2021), and the number of cases growled up to 48 cases in weeks 23 and 24 and declined therefore (Figure 1). The beginning of the outbreak was detected in a car repair shop located in a district of Pamplona. These cases could be related to another more significant outbreak of variant B.1.575.2, which was found in a mosque. Since these first cases, the variant has spread throughout Pamplona and its surroundings without affecting the rest of Navarra. The median age of patients was 33±17 years old, 46.2% women, 53.8% men and approximately 50% Arab origin. Eighty-two (43.1%) patients acquired the infection at domiciliary ambit, the most common cause. Only 14 (7.1%) acquired it at the workplace. One hundred and fifty-six (79.7%) patients showed symptoms, and only four (2.2%) were admitted to hospitals, but none was a severe case. Six (3.3%) of patients have been fully vaccinated for COVID-19 and 35 (19.2%) had received any vaccine dose.

**Figure 1.**
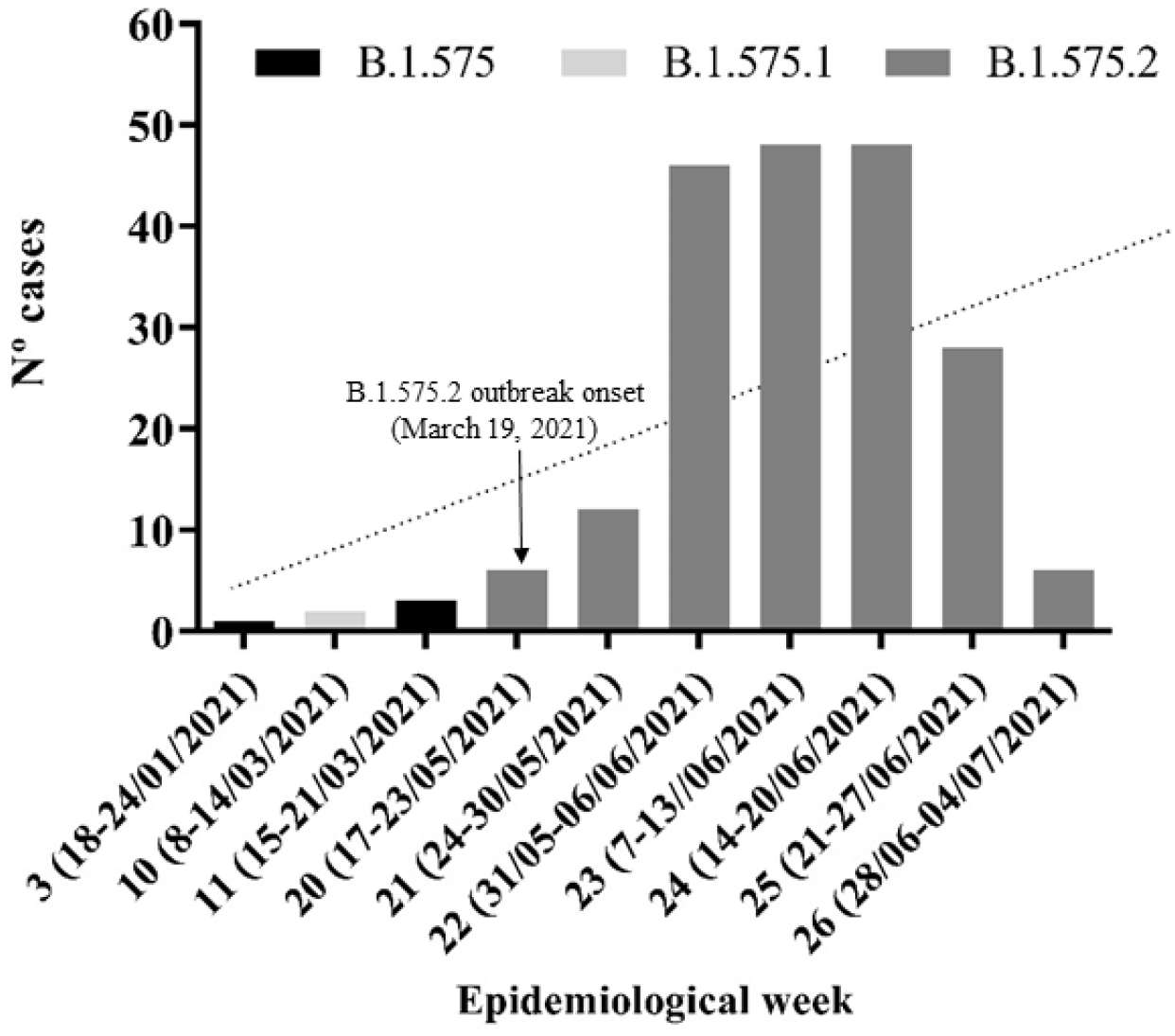
Timeline of SARS-CoV-2 B.1.575, B1.575.1 and B.1.575.2 linages emergence in Pamplona between January and June 2021.

To know the distribution of SARS-CoV-2 B.1.575 lineage, we searched in the GISAID EpiCoV and PANGO lineages databases. From May to July, the lineage and sub-lineages of B.1.575 have increased exponentially in different countries. The B.1.575 lineage was predominant in the United States of America (USA) (90%), while the B.1.575.1 and B.1.575.2 sub-lineages dominated Spain with 86% and 92%, respectively.

The B.1.575.2 sub-lineage was predominant in Navarra since 99.5% (194/197) of the cases registered in the GISAID EpiCoV database were identified in this region. By contrast, we did not identify any genomes with B.1.575 and B.1.575.1 lineage carrying the E484K mutation.

## Conclusions

In this study, we observed the emergence of a lineage B.1.575.2 to acquire the spike E484K mutation circulating in Pamplona associated an outbreak. The new lineage displayed a low prevalence (4.10%) among SARS-CoV-2 genomes analyzed between March 23, 2020, and June 30, 2021. Still, it is already dispersed in our city and comprises 97% of the B.1.575 sequences detected in that period. The E484K mutation is considered one of the most important substitutions associated with reduced antibody neutralization potency and efficacy of the SARS-CoV-2 vaccine (12-14). The E484K mutation has been identified in SARS-CoV-2 variants considered VOC such as B.1.351, P.1 and B.1.1.7+E484K and in VOI variants such as B.1.525, B.1.620, and B1.621 among others (1-3), so the presence of this mutation should be supervised and monitored.

Screening PCR is a useful tool for detecting mutations, mainly because of its rapidity. Future identifications with this method could include new mutations characteristic of the lineage could serve as a rapid method of variant identification. However, whole genome sequencing remains the gold standard technique for pandemic control. This brief report emphasizes the importance of exhaustive surveillance for circulating variants of the SARS-CoV-2 virus to reduce community transmission and prevent the emergence of more transmissible variants that could further increase the severity of the epidemic in the country.

## Data Availability

All genomes generated in this work were deposited in the GISAID EpiCoV database (http://gisaid.org).

## Conflict of interest

The authors declare no conflict of interest.

## Funding

This work was supported by the Horizon 2020 program of the European Commission (I-MOVE-COVID-19, grant agreement No 101003673) and the Carlos III Institute of Health with the European Regional Development Fund (COV20/00542).

## Acknowledgements

We would like to thank the GISAID EpiCoV database and all contributing researchers for making the genomes publicly available.

## Author contributions

CTS, AM, MEP, MFH AN, CE conceived and designed the study. GRO was responsible for the whole genome sequencing interpretation. PSS, PLM, JC provided epidemiology data. CTS wrote the manuscript, and all authors critically revised the manuscript. All authors approved the final version of the manuscript and were accountable for its accuracy.

## Notes

### Competing Interest Statement

The authors have declared no competing interest.

### Author Declarations

The Ethical Committee approved the study protocol for Clinical Research of Navarra, PI_2020/96

